# Occurrence of human infection with *Salmonella* Typhi in sub-Saharan Africa

**DOI:** 10.1101/2023.09.22.23295884

**Authors:** Jong-Hoon Kim, Prerana Parajulee, Thuy Tien Nguyen, Shreeya Wasunkar, Vittal Mogasale, Se Eun Park, Ursula Panzner, Ondari D. Mogeni, Justin Im, Florian Marks

## Abstract

Typhoid fever, caused by *Salmonella enterica* serovar Typhi, results in over 1.2 million cases and 29 thousand deaths annually from sub-Saharan Africa. Combating this disease requires various intervention approaches, such as typhoid conjugate vaccines and improving water, sanitation, and hygiene. Enhancing the effectiveness of these strategies necessitates a deeper understanding of the variation of the typhoid fever across the target region. Although the magnitude and variation of typhoid fever at the country level have been studied globally, sub-national variation remains underexplored. To address this gap, we collected data from 265 published reports on typhoid fever occurrences in sub-Saharan Africa between January 2000 and December 2020. The dataset includes information on the year and geographical location of observation, diagnostic tests used, and the type of studies in which typhoid fever was reported. By analyzing this dataset, we can gain insights into the sub-national heterogeneity of typhoid fever’s burden in the region. This knowledge will be instrumental in designing more effective intervention strategies to combat the disease.

## Background & Summary

Typhoid fever is a systemic bacterial infection caused by *Salmonella enterica* serovar Typhi (*Salmonella* Typhi or *S*. Typhi). Patients usually present sustained fever (39-40°C) and other symptoms include weakness, stomach pain, headache, diarrhea or constipation, cough, and loss of appetite. Severe forms of illness include illeal perforation, which can lead to death. *S*. Typhi is transmitted via fecally-contaminated food and water and the majority of typhoid fever incidence is known to occur in low- and middle-income countries (LMIC)^1-6^.

Globally, typhoid fever causes estimated 12 million cases and 130 thousand deaths according a recent modelling study in which incidence rate data come from population-based longitudinal surveillance studies conducted at 22 sites in 14 different countries between 1978 and 2017.^1^ Earlier estimates of the global burden of typhoid fever were based on a relatively simplistic approach of extrapolating the incidence rates observed in one setting to the entire country or to neighbouring countries where data is unavailable^2–4^. However, the surveillance is often conducted at sites where the disease incidence has already been reported and therefore would not necessarily well represent the country^5^. Later studies used sophisticated modelling techniques to adjust observed values using the distribution of geospatial variables that potentially affect the transmission of typhoid fever rather than simple extrapolaton^1,6^.

While they clearly show variation at the country level, existing studies fail to emphasize that the burden of typhoid fever also shows significant sub-national variation for each country. Outbreaks often show district-level variation of typhoid incidence^7–9^ and country-level surveys show sub-national heterogeneity of incidence^10–13^. Understanding idiosyncratic behaviour of typhoid transmission between communities will be critical for a country to implement intervention programs such as campaign vaccination against endemic or epidemic typhoid fever more efficiently and effectively. Efficiency and effectiveness of an intervention program can improve, for instance, if the interventions are targeted on high-burden areas. Identifying those high-burden areas will depend on how well we understand the spatial variation of the burden of typhoid fever across the target country.

Although population-based prospective studies serve as the basis for existing estimates of country-level and global burden of typhoid fever, report of sporadic cases and outbreaks provide wider spatial coverage and hold information on sub-national variation of occurrence of typhoid fever^14^. In this study, we provide the data set on the occurrence of typhoid fever extracted from peer-reviewed literature. We focus on sub-Saharan Africa, where systematic surveillance has shown that the burden is substantial^1,2,5,6^. The dataset we report provides information on the occurrence of typhoid fever at the hospital, or district or higher sub-national administrative units. These datasets provide opportunities to better understand sub-national variation of the occurrence of typhoid fever.

## Methods

### Search and Data Sources

Data on the occurrence of typhoid fever were extracted from peer-reviewed research articles indexed in PubMed or Embase published between January 1, 2000 and October 31, 2020. Systematic review of the literature followed a standard procedure and the overview of the procedure in a Preferred Reporting Items for Systematic Reviews and Meta-Analyses (PRISMA) flow diagram (**Figure 1**). The search term used for querying PubMed and Embase was designed to capture different uses of typhoid fever in the literature such as typhoid fever, *Salmonella* typhi, *S*. Typhi, or enteric fever and limit to the countries from sub-Saharan Africa. The exact search terms appear in the Supplement.

**Figure 1.**
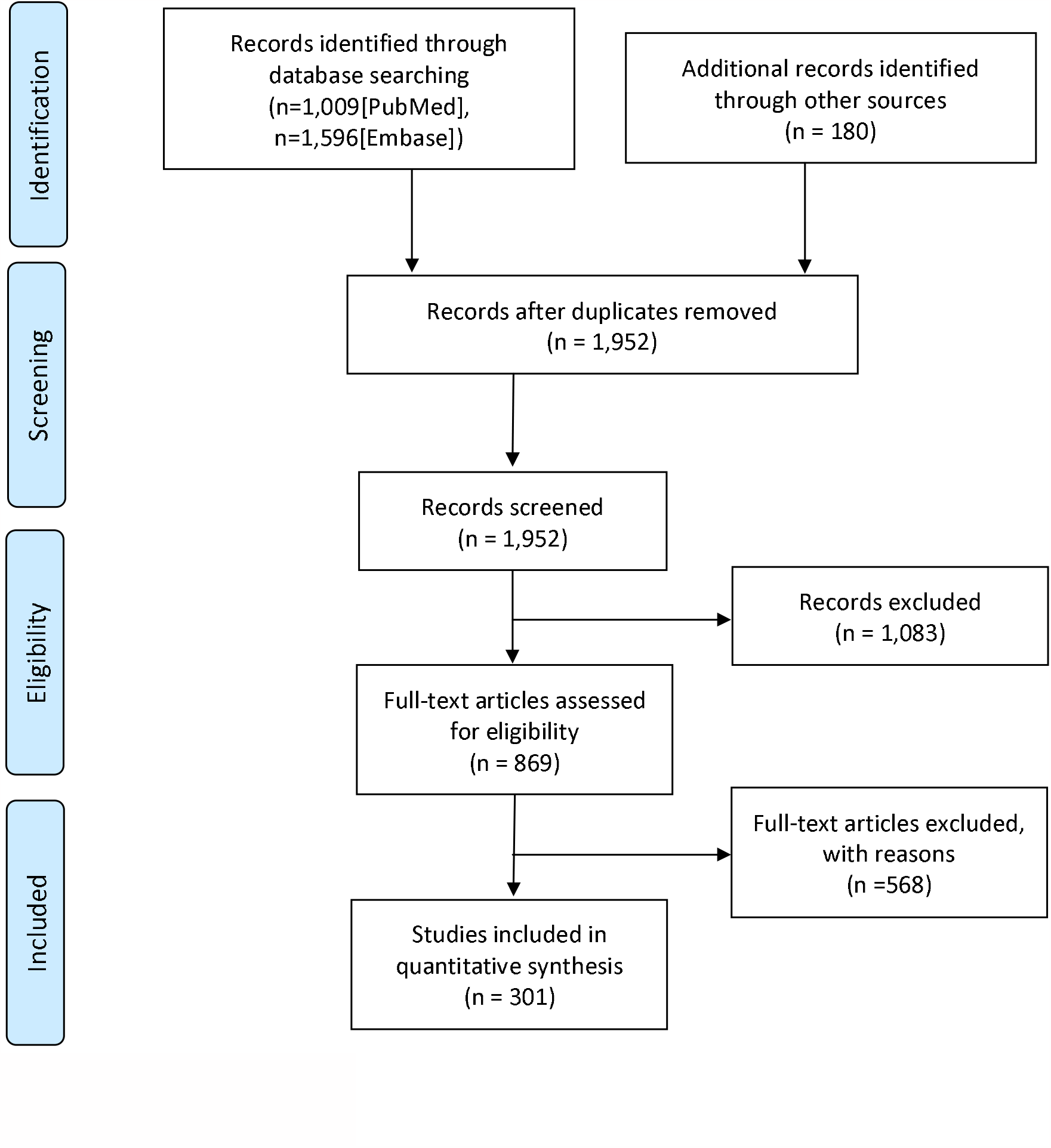
PRISMA flow diagram

### Eligibility Criteria and Study Selection

A study, regardless of study types, was eligible for inclusion in this analysis if it clearly reports date and location of occurrence as well as diagnostic method used for confirmation of the occurrence of human typhoid cases. We included reports of polymerase chain reaction (PCR) or culture-confirmed typhoid fever where *S*. Typhi was isolated from normally sterile sites such as blood, urine, bone marrow, or cerebrospinal fluid. We also included studies that confirm typhoid fever through serologic tests (e.g., Widal test or enzyme-linked immunosorbent assay [ELISA]) or suspected clinically (e.g., ileal perforation) while acknowledging that those case definitions are less reliable compared with culture or PCR confirmation. We excluded the studies that are based on the analyses of existing data and do not report novel occurrence of typhoid fever in humans. For instance, some articles report results of experiments using the existing isolates (e.g., susceptibility to antimicrobials or other medicinal plants) and were therefore excluded. Other studies fail to provide serovars while reporting infection Salmonella.

### Data Extraction, Study Variables, and Analytic Approach

Two authors (J-HK and PP) reviewed the literature and extracted the data. Where there was discordance among the two reviewers, the first author decided after discussions. Extracted data included year of observation, location (smallest sub-national area possible), diagnostic method, and the number of typhoid cases reported. In reports of typhoid cases from observations that span multiple years without further details broken down by year, we assumed that at least 1 episode of typhoid fever case occurred each year. We defined a typhoid fever occurrence as a report of at least 1 episode of typhoid fever excluding any duplicate reports from the same cohort. For imported cases, we assumed that the typhoid episode occurred in the country in which the infection occurred according to the report. If the imported cases led to local transmission, we assumed that typhoid occurred in both the country of infection origin and the country of report. An outbreak was defined based on the definition used in the report.

## Data Records

Occurrence data were stored in an Excel spreadsheet where each row indicates an occurrence event of typhoid fever for specific year and location. Some occurrence events reported from a larger region may overlap those from smaller sub-regions within a region. Columns indicate data source (e.g., title of the article), years of observation and report, diagnostic method, number of cases reported, sub-national region (up to the smallest units available), study type, longitude and latitude which were acquired through Google Map. Locations may refer to the arbitrary central point of the region (e.g., neighbourhoods, village) or the location of a healthcare facility. Study types were categorized into review of hospital records, case report, outbreak reports, and longitudinal studies. The dataset is available at the GitHub repository of the first author.^15^

## Technical Validation

We provide an overview of the included studies by country and year of publication. Also, we provide more details on the diagnostic methods used to confirm the infection, types of studies in which typhoid fever were reported, year, and sub-national location of occurrence of typhoid fever.

### Frequency of studies by year and country

Occurrence of confirmed typhoid fever were reported in 31 countries from Jan 2000 to Dec 2020 with overall reporting frequency increasing over time (**Figure 2A**). The number of reports varies by study and year of publication (**Figure 2B**) and by year in which the study began (**Figure 2C**) considerably. Nigeria consistently reported the highest number of reports on the occurrence of typhoid fever over the period. For countries like Angola, Benin, Burundi, Guinea-Bissau, Liberia, Madagascar, Niger, Uganda, Zambia and Zimbabwe, typhoid fever has been reported after 2010 considering the period from 2000 to 2020.

**Figure 2.**
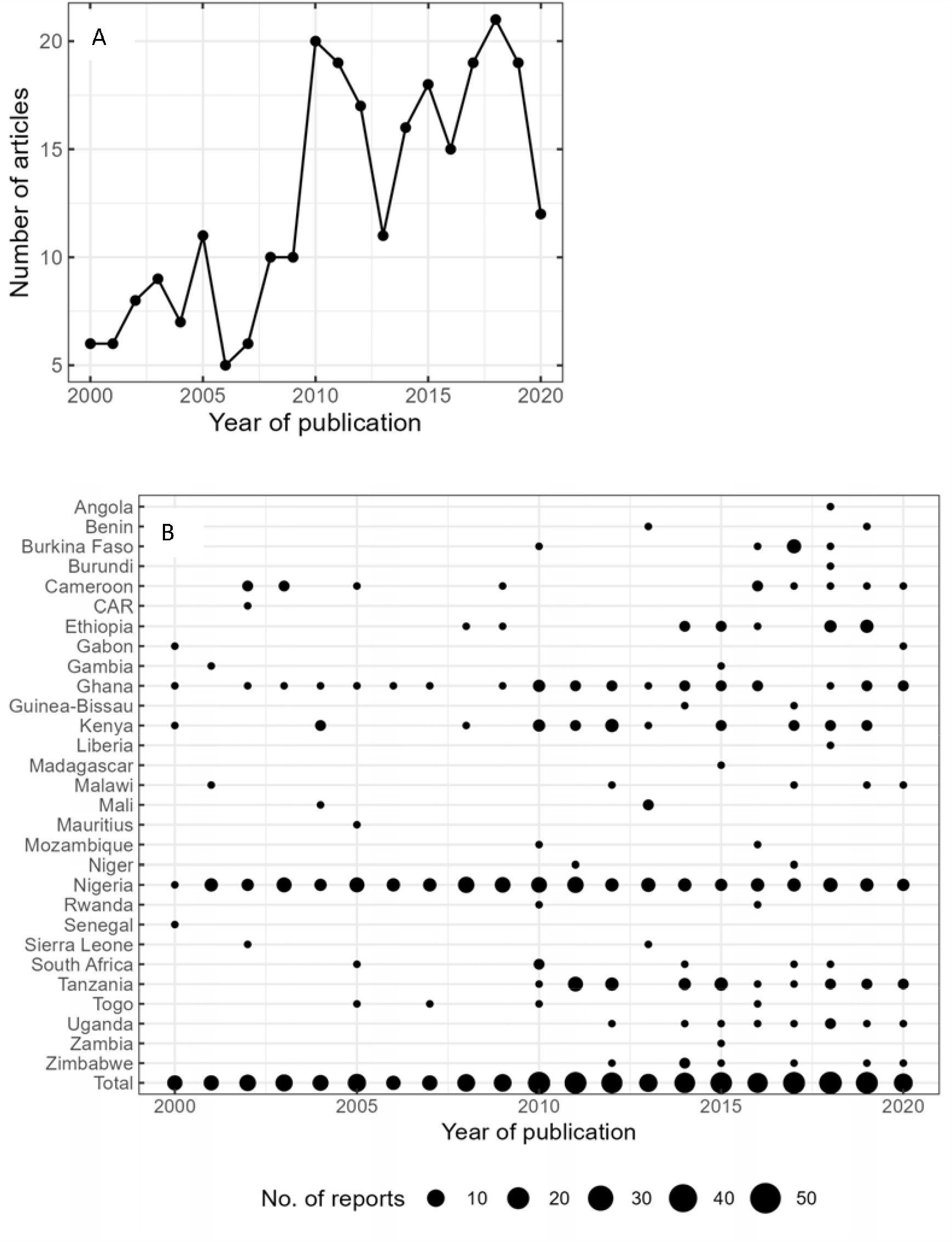
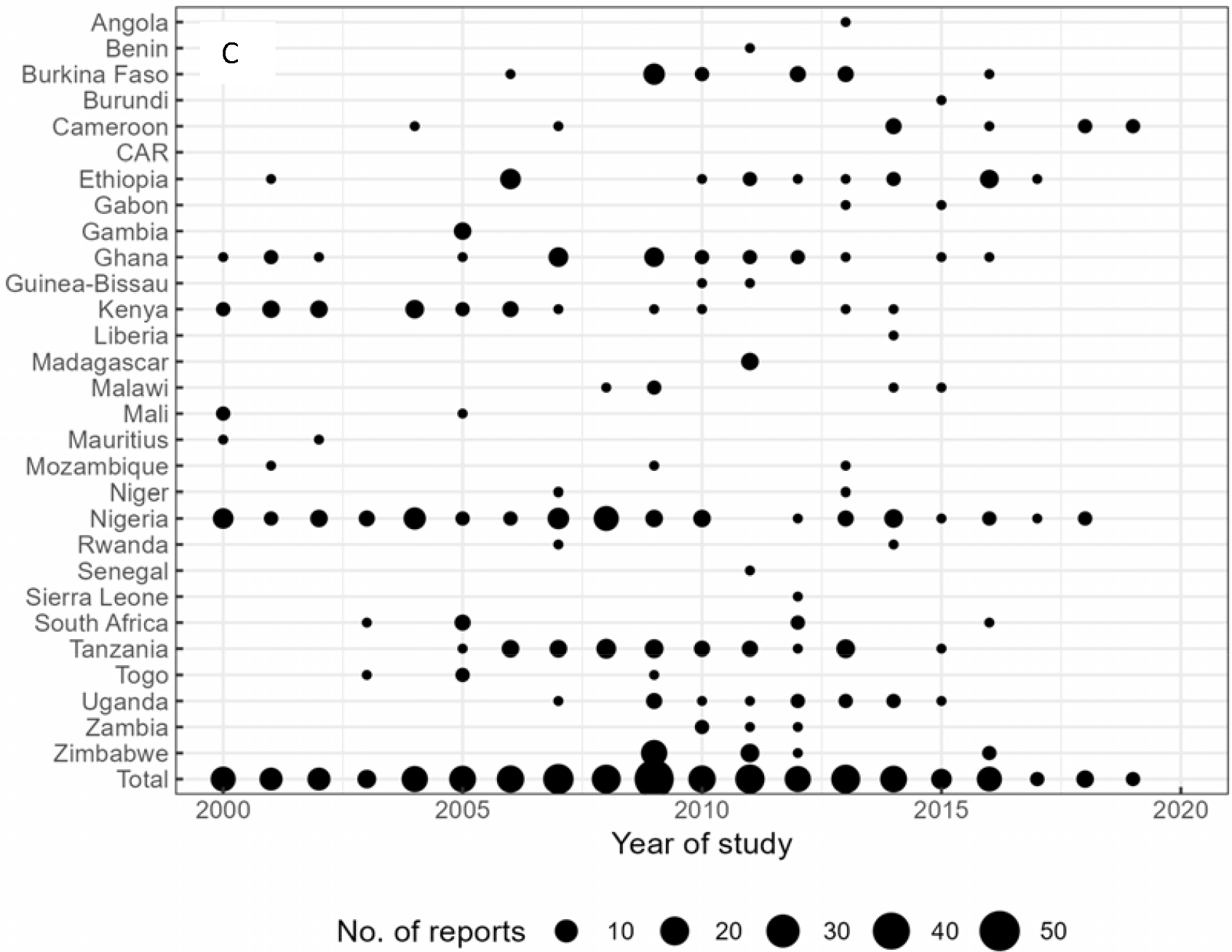
Number of included reports by year and country. The overall number of reports of typhoid occurrence in sub-Saharan Africa increases over time (A) with the considerable variation by year of publication and country (B). The number of studies showed substantial variation also by country and the year in which the study began. CAR = Central African Republic

### Diagnostic methods

We categorized the studies based on the diagnostic method employed to identify typhoid fever (*Table 1*). In instances where a single study utilized multiple diagnostic methods (e.g., clinical criteria, blood culture, and bone marrow sample culture), we prioritized the method with the higher performance. Specifically, if the study included the method such as the culture of bone marrow or cerebrospinal fluid samples, which is recognized as the highest sensitivity, we selected this method as the representative method of the study. Similarly, a study incorporated blood culture but did not included methods like the bone marrow culture, we chose the blood culture as the representative diagnostic method. If the methods included other culture methods, we chose the methods in the order of culture of urine samples, culture of stool samples, and culture of other body fluids. Other studies did not specifically mention the samples used to culture but implied the culturing method (e.g., “bacterial isolation” or “microbiologically confirmed”). In this case, we created a category called “Culture”. For studies whose diagnostic methods do not include culturing method, we chose the method in an order of Rapid tests, Widal, biochemical tests, and clinical symptoms. While we sought to categories by the precision of the tests, this category maybe somewhat arbitrary. The reader can access the original terms that indicated multiple methods used for each study.

**Table 1.** Frequency of studies by diagnostic methods. CSF = Cerebrospinal fluid.

| Diagnostic method | Frequency [n (%)] | Samples of actual terms used in the literature |
| --- | --- | --- |
| Culture of blood samples | 99 (37.4) | "urine, blood, or stool culture", "blood or stool culture, or Widal-Felix", "blood/urine/stool culture", etc. |
| Clinical symptoms | 71 (26.8) | "typhoid perforation", "intestinal perforation", "clinical diagnosis", "digestive perforation", "typhoid gall bladder perforation", "cholecystitis", "clinical", "hospital record", "ileal perforation", "multiple jejunal perforations", "perforated enteritis", "peritonitis", "small bowel perforation", "enterocutaneous fistula", "septicaemia", "case files", "cases reported", "death records", "typhoid fever", etc. |
| Widal test | 36 (13.6) | "Widal test", "agglutination and titration (> 4-fold)", "Widal-Felix", "Widal and Weilfelix direct card agglutination tests (DCAT)", "Tube agglutination test", etc. |
| Culture of stool samples | 17 (6.4) | "stool culture", etc. |
| Culture | 14 (5.3) | "Isolation of Salmonella typhi", "culture", "isolated", "bacterial isolation", "microbiologically confirmed", "laboratory confirmed", "confirmed", etc. |
| Unclear | 11 (4.2) | "Survey questionnaire", "surveillance data", "National Health Laboratory Services data", "standard methods with blood samples", "travel or tropical medicine clinics record", "history based on self report", "questionnaire", |
|  |  | "report at medical record", "self reported" |
| Culture of CSF samples | 5 (1.9) | "cerebrospinal fluid culture", "spinal fluid culture",<br>"isolation from CSF, blood, or stool", etc. |
| Autopsy | 3 (1.1) | "post mortem report", "autopsy", "coroner's autopsies" |
| Culture of bone marrow samples | 3 (1.1) | "blood, stool or bone marrow culture", "blood and bone marrow cultures", etc. |
| Rapid test | 2 (0.8) | "rapid typhoid test", "Blood-RDT", "rapid diagnostic test",<br>"RDT", etc. |
| Culture of other body fluids | 1 (0.4) | "aspirates culture", "culture of pus", "wound culture",<br>"wound swab culture", "Ear and throat culture", "Sputum culture", "rectal swab specimens examined bacteriologically", etc. |
| Biochemical test | 1 (0.4) | "biochemical", "biochemical testing of urine sample",<br>"biochemical tests of environmental samples", etc. |
| Culture of environmental samples | 1 (0.4) | "environmental sample culture", "S. typhi isolated from water" |
| Culture of urine samples | 1 (0.4) | "urine culture" |

According to this categorization, studies based on culturing blood samples were most frequent (*n*=99, 37.4%). (**Table 1**) while still numerous studies relied on less reliable methods, which include clinical signs (e.g., illeal perforation) (*n*=71, 26.8%) or Widal test (or agglutination) (*n*=36, 13.6%).

### Study types

Study design was diverse and included case reports, outbreak investigation, cross-sectional studies, retrospective studies (i.e., review of hospital records), and prospective studies including multi-year longitudinal surveillance studies. While population-based longitudinal surveillance studies can provide incidence rates of typhoid fever, they are few (n = 5) and were conducted around 15 sub-national areas of 10 countries (**Table 2**). On the other hand, there are other studies that report occurrence of typhoid fever and wider spatial coverage in the dataset. Prospective studies including clinical trials were most common (n=212), followed by retrospective (n=144), cross-sectional (n=48), outbreak investigation (n=26), case report (n=26), and case-control (n=13) studies.

**Table 2.** Population-based longitudinal surveillance of typhoid fever in sub-Saharan Africa since 2000.

| Country | Sub-national region | Surveillance period | Ref |
| --- | --- | --- | --- |
| Burkina Faso | Nanoro, Boulkiemde | 2013-2014 | <sup>16</sup> |
| Burkina Faso | Nioko II, Polesgo | 2012-2013 | <sup>5</sup> |
| Ethiopia | Butajira | 2012-2014 | <sup>5</sup> |
| Ghana | Asante Akim North | 2007-2009 | <sup>17</sup> |
| Ghana | Asante Akim North | 2010-2012 | <sup>5</sup> |
| Guinea-Bissau | Bandim | 2011-2013 | <sup>5</sup> |
| Kenya | Lwak, Kibera | 2006-2009 | <sup>13</sup> |
| Kenya | Kibera | 2012-2013 | <sup>5</sup> |
| Madagascar | Imerintsiaosika, Isotry | 2011-2013 | <sup>5</sup> |
| Senegal | Pikine | 2011-2013 | <sup>5</sup> |
| South Africa | Pietermaritzburg | 2012-2014 | <sup>5</sup> |
| Sudan | East Wad Medani | 2012-2013 | <sup>5</sup> |
| Tanzania | Pemba Island | 2010-2012 | <sup>18</sup> |
| Tanzania | Moshi urban, Moshi rural | 2011-2013 | <sup>5</sup> |

### Geographical locations

Occurrence data cover entire continent although occurrences are more frequent in East and West Africa. Top three countries in terms of the number of unique 20 km by 20 km grids where the occurrence was reported were Nigeria (n=85), Kenya (n=24), and Tanzania (n=23) (**Figure 3A-D**). Sub-national locations in the dataset includes household, primary clinic, tertiary hospitals, district, or provinces. Except for the household locations (n=180), occurrence of typhoid fever has been reported in small number of large hospitals and cities with substantial variation across countries. In Angola, National Institute of Public Health of Angola located in the Capital city, Luanda is the only place in which blood culture-confirmed typhoid fever was reported. On the other hand, more than 55 health centers and hospitals spread out through the country.

**Figure 3.**
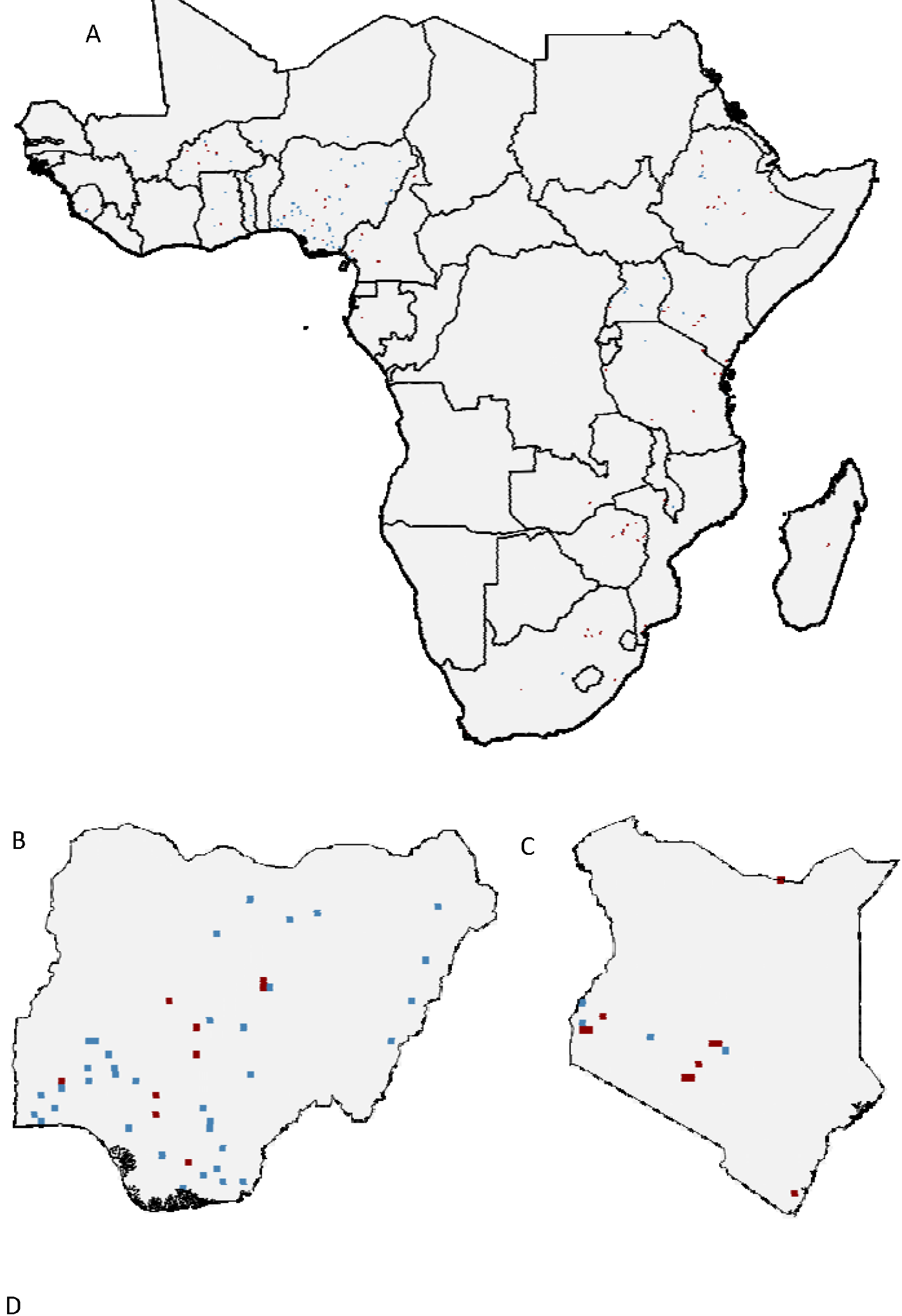
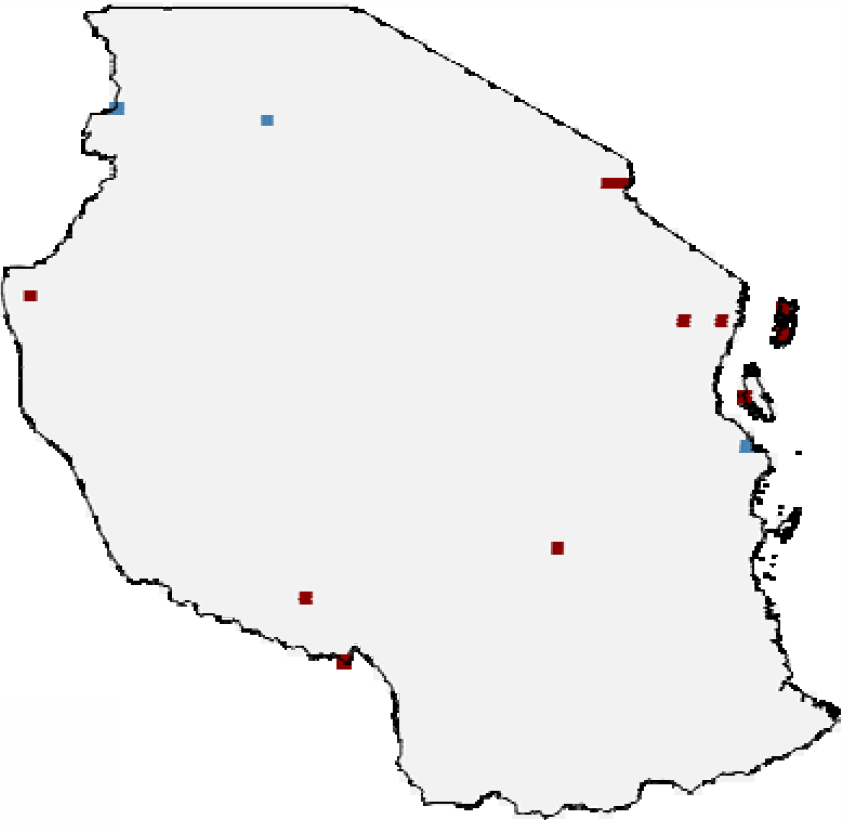
Geographical locations of occurrence of typhoid fever. The map has 20 km by 20 km resolution. Red and blue grids represent location of typhoid fever confirmed through culture and other methods, respectively in Africa (A). (B), (C) and (D) represent Nigeria, Kenya, and Tanzania, respectively, in which typhoid occurrences were most frequently reported.

## Data Availability

All data produced are available online at https://github.com/kimfinale/typhoid_occurrence.

## Code Availability

All the codes used to generate the figures and tables are available in the GitHub repository ^15^

## Author contributions

J-HK designed the study, performed the literature search, reviewed the articles, extracted the data, and wrote the manuscript. PP reviewed the articles, extracted data, and reviewed the manuscript. All authors reviewed the final version of the manuscript and approved its submission.

