## Supplementary material for "Occurrence of human infection with *Salmonella* Typhi in sub-Saharan Africa": Supplmentary Material

1. PubMed

("Typhoid Fever"[Mesh] OR "Salmonella typhi"[Mesh] OR “typhoid,”[tw] OR “S. Typhi,” [tw] OR “Salmonella Typhi,” [tw] OR “enteric fever”[tw])

AND

hasabstract

AND

("Africa South of the Sahara"[Mesh] OR "sub-saharan Africa"[Tiab] OR "subsaharan Africa"[Tiab] OR "Sub Saharan Africa"[Tiab] OR "Africa, Central"[Mesh] OR "Central Africa"[Tiab] OR "Cameroon"[Mesh] OR "Cameroon"[Tiab] OR "Central African Republic"[Mesh] OR "Central African Republic"[Tiab] OR "Chad"[Mesh] OR "Chad"[Tiab] OR "Congo"[Mesh] OR "Congo"[Tiab] OR "Democratic Republic of the Congo"[Mesh] OR "Democratic Republic of the Congo"[Tiab] OR "Equatorial Guinea"[Mesh] OR "Equatorial Guinea"[Tiab] OR "Gabon"[Mesh] OR "Gabon"[Tiab] OR "Sao Tome and Principe"[Mesh] OR "Sao Tome and Principe"[Tiab] OR "Africa, Eastern"[Mesh] OR "Eastern Africa"[Tiab] OR "East Africa"[Tiab] OR "Burundi"[Mesh] OR "Burundi"[Tiab] OR "Djibouti"[Mesh] OR "Djibouti"[Tiab] OR "Eritrea"[Mesh] OR "Eritrea"[Tiab] OR "Ethiopia"[Mesh] OR "Ethiopia" [Tiab] OR "Kenya"[Mesh] OR "Kenya" [Tiab] OR "Rwanda"[Mesh] OR "Rwanda" [Tiab] OR "Somalia"[Mesh] OR "Somalia"[Tiab] OR "South Sudan"[Mesh] OR "South Sudan"[Tiab] OR "Sudan"[Mesh] OR "Sudan"[Tiab] OR "Tanzania"[Mesh] OR "Tanzania"[Tiab] OR "Uganda"[Mesh] OR "Uganda"[Tiab] OR "Africa, Southern"[Mesh] OR "Southern Africa"[Tiab] OR "South Africa"[Tiab] OR "Angola"[Mesh] OR "Angola"[Tiab] OR "Botswana"[Mesh] OR "Botswana"[Tiab] OR "Lesotho"[Mesh] OR "Lesotho"[Tiab] OR "Malawi"[Mesh] OR "Malawi"[Tiab] OR "Mozambique"[Mesh] OR "Mozambique"[Tiab] OR "Namibia"[Mesh] OR "Namibia"[Tiab] OR "South Africa"[Mesh] OR "South Africa"[Tiab] OR "Swaziland"[Mesh] OR "Swaziland"[Tiab] OR "Zambia"[Mesh] OR "Zambia"[Tiab] OR "Zimbabwe"[Mesh] OR "Zimbabwe"[Tiab] OR "Africa, Western"[Mesh] OR "Western Africa"[Tiab] OR "West Africa"[Tiab] OR "Benin"[Mesh] OR "Benin"[Tiab] OR "Burkina Faso"[Mesh] OR "Burkina Faso"[Tiab] OR "Cabo Verde"[Mesh] OR "Cabo Verde"[Tiab] OR "Cote d'Ivoire"[Mesh] OR "Cote d'Ivoire"[Tiab] OR "Gambia"[Mesh] OR "Gambia"[Tiab] OR "Ghana"[Mesh] OR "Ghana"[Tiab] OR "Guinea"[Mesh] OR "Guinea"[Tiab] OR "Guinea-Bissau"[Mesh] OR "Guinea-Bissau"[Tiab] OR "Liberia"[Mesh] OR "Liberia"[Tiab] OR "Mali"[Mesh] OR "Mali"[Tiab] OR "Mauritania"[Mesh] OR "Mauritania"[Tiab] OR "Niger"[Mesh] OR "Niger"[Tiab] OR "Nigeria"[Mesh] OR "Nigeria"[Tiab] OR "Senegal"[Mesh] OR "Senegal"[Tiab] OR "Sierra Leone"[Mesh] OR "Sierra Leone"[Tiab] OR "Togo"[Mesh] OR "Togo"[Tiab] OR "Comoros"[Mesh] OR "Comoros"[Tiab] OR "Mayotte"[Tiab] OR "Madagascar"[Mesh] OR "Madagascar"[Tiab] OR "Sahel"[Tiab])

2. Embase

(‘typhoid’:ti,ab OR ‘S. Typhi’:ti,ab OR ‘Salmonella Typhi’:ti,ab OR ‘enteric fever’:ti,ab)

AND

'Africa south of the Sahara'/exp OR ‘Sub-Saharan Africa’:ti,ab OR ‘Subsaharan Africa’:ti,ab OR ‘Sub Saharan Africa’:ti,ab OR 'Central Africa'/exp OR 'Central Africa':ti,ab OR 'Cameroon'/exp OR 'Cameroon':ti,ab OR 'Central African Republic'/exp OR 'Central African Republic':ti,ab OR 'Chad'/exp OR 'Chad':ti,ab OR 'Congo'/exp OR 'Congo':ti,ab OR 'Democratic Republic Congo'/exp OR 'Democratic Republic Congo':ti,ab OR 'Equatorial Guinea'/exp OR 'Equatorial Guinea':ti,ab OR 'Gabon'/exp OR 'Gabon':ti,ab OR 'Sao Tome and Principe'/exp OR 'Sao Tome and Principe':ti,ab OR ‘Eastern Africa’:ti,ab OR ‘East Africa’:ti,ab OR 'Burundi'/exp OR 'Burundi':ti,ab OR 'Djibouti'/exp OR 'Djibouti':ti,ab OR 'Eritrea'/exp OR 'Eritrea':ti,ab OR 'Ethiopia'/exp OR 'Ethiopia':ti,ab OR 'Kenya'/exp OR 'Kenya':ti,ab OR 'Rwanda'/exp OR 'Rwanda':ti,ab OR 'Somalia'/exp OR 'Somalia':ti,ab OR 'South Sudan'/exp OR 'South Sudan':ti,ab OR 'Sudan'/exp OR 'Sudan':ti,ab OR 'Tanzania'/exp OR 'Tanzania':ti,ab OR 'Uganda'/exp OR 'Uganda':ti,ab OR ‘Southern Africa’:ti,ab OR ‘South Africa’:ti,ab OR 'Angola'/exp OR 'Angola':ti,ab OR 'Botswana'/exp OR 'Botswana':ti,ab OR 'Lesotho'/exp OR 'Lesotho':ti,ab OR 'Malawi'/exp OR 'Malawi':ti,ab OR 'Mozambique'/exp OR 'Mozambique':ti,ab OR 'Namibia'/exp OR 'Namibia':ti,ab OR 'South Africa'/exp OR 'South Africa':ti,ab OR 'Swaziland'/exp OR 'Swaziland':ti,ab OR 'Zambia'/exp OR 'Zambia':ti,ab OR 'Zimbabwe'/exp OR 'Zimbabwe':ti,ab OR ‘Western Africa’:ti,ab OR ‘West Africa’:ti,ab OR 'Benin'/exp OR 'Benin':ti,ab OR 'Burkina Faso'/exp OR 'Burkina Faso':ti,ab OR 'Cape Verde'/exp OR 'Cape Verde':ti,ab OR 'Cote d`Ivoire'/exp OR 'Cote d`Ivoire':ti,ab OR 'Gambia'/exp OR 'Gambia':ti,ab OR 'Ghana'/exp OR 'Ghana':ti,ab OR 'Guinea'/exp OR 'Guinea':ti,ab OR 'Guinea-Bissau'/exp OR 'Guinea-Bissau':ti,ab OR 'Liberia'/exp OR 'Liberia':ti,ab OR 'Mali'/exp OR 'Mali':ti,ab OR 'Mauritania'/exp OR 'Mauritania':ti,ab OR 'Niger'/exp OR 'Niger':ti,ab OR 'Nigeria'/exp OR 'Nigeria':ti,ab OR 'Senegal'/exp OR 'Senegal':ti,ab OR 'Sierra Leone'/exp OR 'Sierra Leone':ti,ab OR 'Togo'/exp OR 'Togo':ti,ab OR 'Comoros'/exp OR 'Comoros':ti,ab OR 'Mayotte'/exp OR 'Mayotte':ti,ab OR 'Madagascar'/exp OR 'Madagascar':ti,ab OR 'Sahel'/exp OR 'Sahel':ti,ab

3. Web of Science

(AB=(((nontyph* OR non-typh* OR group OR typhimurium OR enteritidis OR Heidelberg OR Dublin OR Choleraesuis OR Newport OR Virchow OR Concord OR Brancaster OR freetown OR Infantis OR Isangi) AND (salmonella OR salmonellosis)) OR iNTS)

AND

AB=(bacteremia OR sepsis OR septic* OR Invasive OR bloodstream* OR blood culture OR blood-culture OR hemoculture OR incidence OR epidemiolog* OR burden OR case OR infect* OR prevalen*)

AND

AB=(“Sub-Saharan Africa” OR “Subsaharan Africa” OR “Sub Saharan Africa” OR “Central Africa” OR Cameroon OR “Central African Republic” OR Chad OR Congo OR “Democratic Republic Congo” OR “Equatorial Guinea” OR Gabon OR “Sao Tome and Principe” OR “Eastern Africa” OR “East Africa” OR Burundi OR Djibouti OR Eritrea OR Ethiopia OR Kenya OR Rwanda OR Somalia OR “South Sudan” OR Sudan OR Tanzania OR Uganda OR “Southern Africa” OR “South Africa” OR Angola OR Botswana OR Lesotho OR Malawi OR Mozambique OR Namibia OR Swaziland OR Zambia OR Zimbabwe OR “Western Africa” OR “West Africa” OR Benin OR Burkina Faso OR “Cape Verde” OR “Cote d`Ivoire” OR Gambia OR Ghana OR Guinea OR “Guinea-Bissau” OR Liberia OR Mali OR Mauritania OR Niger OR Nigeria OR Senegal OR “Sierra Leone” OR Togo OR Comoros OR Mayotte OR Madagascar OR Sahel))

OR

(TI=(((nontyph* OR non-typh* OR group OR typhimurium OR enteritidis OR Heidelberg OR Dublin OR Choleraesuis OR Newport OR Virchow OR Concord OR Brancaster OR freetown OR Infantis OR Isangi) AND (salmonella OR salmonellosis)) OR iNTS)

AND

TI=(bacteremia OR sepsis OR septic* OR Invasive OR bloodstream* OR blood culture OR blood-culture OR hemoculture OR incidence OR epidemiolog* OR burden OR case OR infect* OR prevalen*)

AND

TI=(“Sub-Saharan Africa” OR “Subsaharan Africa” OR “Sub Saharan Africa” OR “Central Africa” OR Cameroon OR “Central African Republic” OR Chad OR Congo OR “Democratic Republic Congo” OR “Equatorial Guinea” OR Gabon OR “Sao Tome and Principe” OR “Eastern Africa” OR “East Africa” OR Burundi OR Djibouti OR Eritrea OR Ethiopia OR Kenya OR Rwanda OR Somalia OR “South Sudan” OR Sudan OR Tanzania OR Uganda OR “Southern Africa” OR “South Africa” OR Angola OR Botswana OR Lesotho OR Malawi OR Mozambique OR Namibia OR Swaziland OR Zambia OR Zimbabwe OR “Western Africa” OR “West Africa” OR Benin OR Burkina Faso OR “Cape Verde” OR “Cote d`Ivoire” OR Gambia OR Ghana OR Guinea OR “Guinea-Bissau” OR Liberia OR Mali OR Mauritania OR Niger OR Nigeria OR Senegal OR “Sierra Leone” OR Togo OR Comoros OR Mayotte OR Madagascar OR Sahel))
